# Usage of Mineralocorticoids and Isotonic Crystalloids in Subarachnoid Hemorrhage Patients in the United States

**DOI:** 10.1101/2023.09.28.23296245

**Authors:** Akshitkumar M. Mistry, Janki Naidugari, Jocelyn Craven, Logan Williams, Jonathan Beall, Pooja Khatri, Joseph P. Broderick, Todd W. Rice, Hooman Kamel, William Mack

**Affiliations:** Department of Neurosurgery, University of Louisville, Louisville, KY, USA; School of Medicine, University of Louisville, Louisville, KY, USA; Department of Public Health Sciences, Medical University of South Carolina, Charleston, SC, USA; Departments of Neurology and Rehabilitation Medicine, University of Cincinnati College of Medicine, OH, USA; Department of Medicine, Division of Allergy, Pulmonary, and Critical Care Medicine, Vanderbilt University Medical Center, Nashville, TN, USA; Clinical and Translational Neuroscience Unit, Feil Family Brain and Mind Research Institute and Department of Neurology, Weill Cornell Medicine, New York, NY, USA; Department of Neurosurgery, Keck School of Medicine, University of Southern California, Los Angeles, CA, USA

**Keywords:** Subarachnoid hemorrhage, Fludrocortisone, Critical Care, Mineralocorticoid, Fluids

## Abstract

**Background:** The usage rates of mineralocorticoids (fludrocortisone) to treat hyponatremia and isotonic crystalloids (saline and balanced crystalloids) to maintain intravascular volume in patients with aneurysmal subarachnoid hemorrhage (aSAH) patients across the United States are unknown.

**Methods:** We surveyed National Institute of Neurologic Disorders and Stroke (NINDS) StrokeNet sites, which are mostly large, tertiary, academic centers, and analyzed subarachnoid hemorrhage encounters in the Premier Healthcare Database that is representative of all types of hospitals and captures about 20% of all acute inpatient care in the United States.

**Results:** Although mineralocorticoids are used by 70% of the NINDS StrokeNet sites in aSAH patients, it is used in less than 25% of the aSAH encounters in the Premier Database. Although saline is ubiquitously used, balanced crystalloids are increasingly used for fluid therapy in aSAH patients. Its use in the NINDS StrokeNet sites and the Premier Healthcare Database is 41% and 45%, respectively.

**Conclusions:** The use of mineralocorticoids remains low, and balanced crystalloids are increasingly used as fluid therapy in aSAH patients. The effectiveness of mineralocorticoids and balanced crystalloids in improving outcomes for aSAH patients must be rigorously tested in randomized clinical trials.

## INTRODUCTION

Inpatient and overall mortality of patients with aneurysmal subarachnoid hemorrhage (aSAH) are 20% (1-3) and 40%, respectively (4). The high mortality rate results from life-threatening neurological sequelae and non-neurological complications, like fatal volume loss from kidney-drive natriuresis (cerebral salt wasting) (5-7). Ensuing hyponatremia and polyuria can be exceptional in aSAH patients, requiring treatment of hyponatremia and intravenous fluids to maintain their intravascular volume (8-10).

The 2023 American Heart Association aSAH guidelines recommend that “the use of mineralocorticoids is reasonable to treat natriuresis and hyponatremia” (11). Specifically, fludrocortisone is established to reduce natriuresis, hyponatremia, and polyuria in aSAH patients (9). A meta-analysis of the studies shows that mineralocorticoid use lowers the rate of symptomatic vasospasm in aSAH patients (9).

Maintaining intravascular volume is critical in aSAH patients. There are several options for maintenance intravenous fluid therapy: hypotonic or isotonic crystalloid solutions or colloid solutions. The Neurointensive Care consensus and clinical practice guidelines for stroke (including aSAH) and traumatic brain injury *strongly* recommend using crystalloids and against using colloid fluids (12). Given the importance of tonicity to control cerebral edema, isotonic rather than hypotonic crystalloids are most commonly used for intravenous maintenance fluid therapy.

The usage rates of fludrocortisone and different isotonic crystalloids across the United States are unknown. Efforts to understand the impact of these interventions are better informed with this information. In this short communication, we report the usage rates of fludrocortisone and isotonic crystalloids (saline or balanced crystalloids) from a survey of the National Institute of Neurologic Disorders and Stroke (NINDS) StrokeNet sites (which are mostly large, tertiary, academic centers) and in the Premier Healthcare Database.

## METHODS

### NINDS StrokeNet Site Survey

We surveyed the NINDS StrokeNet sites in May 2023. Within the 27 regional coordinating centers, there are 441 clinical performing sites in the StrokeNet network. The survey was filled out by the PIs at 87 sites (19.7% response rate). The following questions were asked:

- In which type of intensive care unit (ICU) does an SAH patient typically get admitted at your site?
  - Dedicated neuro ICU
  - Other ICU
- Is the critical care of patients with aneurysmal SAH managed by a “closed” team? (i.e., only one team placing orders for critical care management, i.e., “closed ICU”)
  - Yes
  - No
- Are plasma electrolytes assessed daily in SAH patients admitted to ICU?
  - Yes
  - No
- How does your site treat hyponatremia in aneurysmal SAH patients? Check all that apply.
  - Mineralocorticoid (i.e., fludrocortisone or hydrocortisone)
  - Enteral salt supplementation
  - Hypertonic saline
  - Desmopressin (DDVAP)
  - Other (specify)
- Does your site have a protocol for treating hyponatremia that all providers follow?
  - Yes, all providers follow our site-specific protocol for treating hyponatremia.
  - No, there is likely provider-dependent variation.
- What crystalloid is used for intravenous fluid therapy in SAH patients at your site? Check all that apply.
  - Saline (0.9% NaCl)
  - Balanced crystalloids (Plasma-Lyte or lactated Ringer’s)
  - Other (specify)

### Premier Healthcare Database

We analyzed the service-level, all-payer Premier® Healthcare Database (PINC AI™ / Premier Inc., Charlotte, NC) of electronic health records and hospital billing data from 2010 to 2020. It is a population-based research database with information from over 900 United States hospitals and health systems. It includes real-world data from at least 45 states in the United States and the District of Columbia, and it is estimated to capture approximately 20% of all acute inpatient care in the United States. It is arguably representative with respect to hospital size, geographic region, location (urban/rural) as well as profit, governmental, and teaching status. Among the data, it includes diagnoses and treatments, such as procedures and medications. Thus, we retrieved inpatient encounters from the Premier Healthcare Database using both the International Classification of Diseases (ICD) 9 and 10 diagnoses codes for subarachnoid hemorrhage (430 and I60) and who also had one or more of the following billing CPT codes for a procedure to treat subarachnoid hemorrhage (61624, 75894, or any code from 61680 to 61711). From these encounters, the total number of encounters with one or more orders of fludrocortisone, saline (normal or 0.9%), and balanced crystalloids (Plasma-Lyte or lactated Ringer’s) was counted.

## RESULTS

### NINDS StrokeNet Site Survey

Three of the 87 NINDS StrokeNet sites that responded to the survey did not answer questions about ICU characteristics (Figure 1). Of the 84 responses, 75 indicated that aSAH patients at their sites were admitted to a dedicated neurological ICU. The ICU is “closed” in 52 sites. In sites where it is “open”, the critical care and neurosurgery teams jointly manage the critical care needs of the patients. All but one site (N=83) that responded assess plasma electrolytes daily.

**Figure 1.**
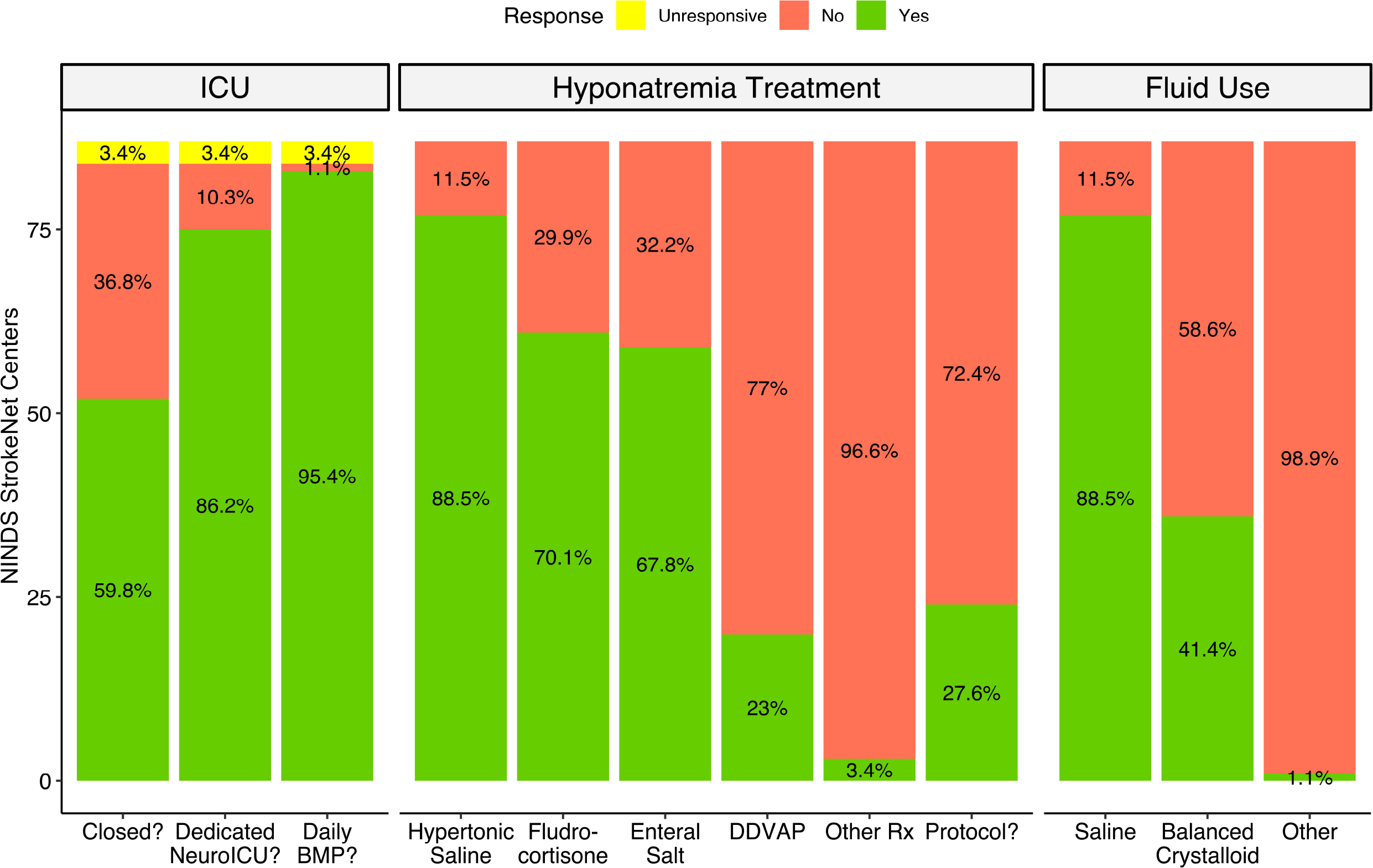
Results of NINDS StrokeNet Sites’ survey, assessing their intensive care unit characteristics, hyponatremia treatments used, and types of fluids used to maintain intravascular volume in aneurysmal subarachnoid hemorrhage patients.

All 87 sites responded to the question of hyponatremia treatment. From the most to least common, treatments used for hyponatremia were hypertonic saline (N=77, 88.5%), mineralocorticoid (N=61, 70.1%), enteral salt supplementation (N=59, 67.8%), and DDVAP (N=20, 23%). Other treatments included fluid restriction (N=5, 5.7%) and urea (N=1). Sixty-three (72.4%) sites do not have a protocol to treat hyponatremia in aSAH patients.

For intravenous fluid therapy, 77 (88.5%) sites use saline, and 36 (41.4%) use balanced crystalloids. Forty-three (49.4%) sites only use saline, and 4 (4.6%) sites only use balanced crystalloids. One site reported using sodium acetate.

### U.S.-Wide Use of Fludrocortisone and Fluids

A total of 20,699 encounters of procedurally treated subarachnoid hemorrhage were retrieved from the Premier Healthcare Database from 2010 to 2020 (Figure 2). The use of fludrocortisone during this time increased gradually but remained less than 25%. For intravenous therapy, saline was used in nearly 100% of the encounters. The use of balanced crystalloids increased from 2010 to 2020, reaching the highest use (45%) in 2020.

**Figure 2.**
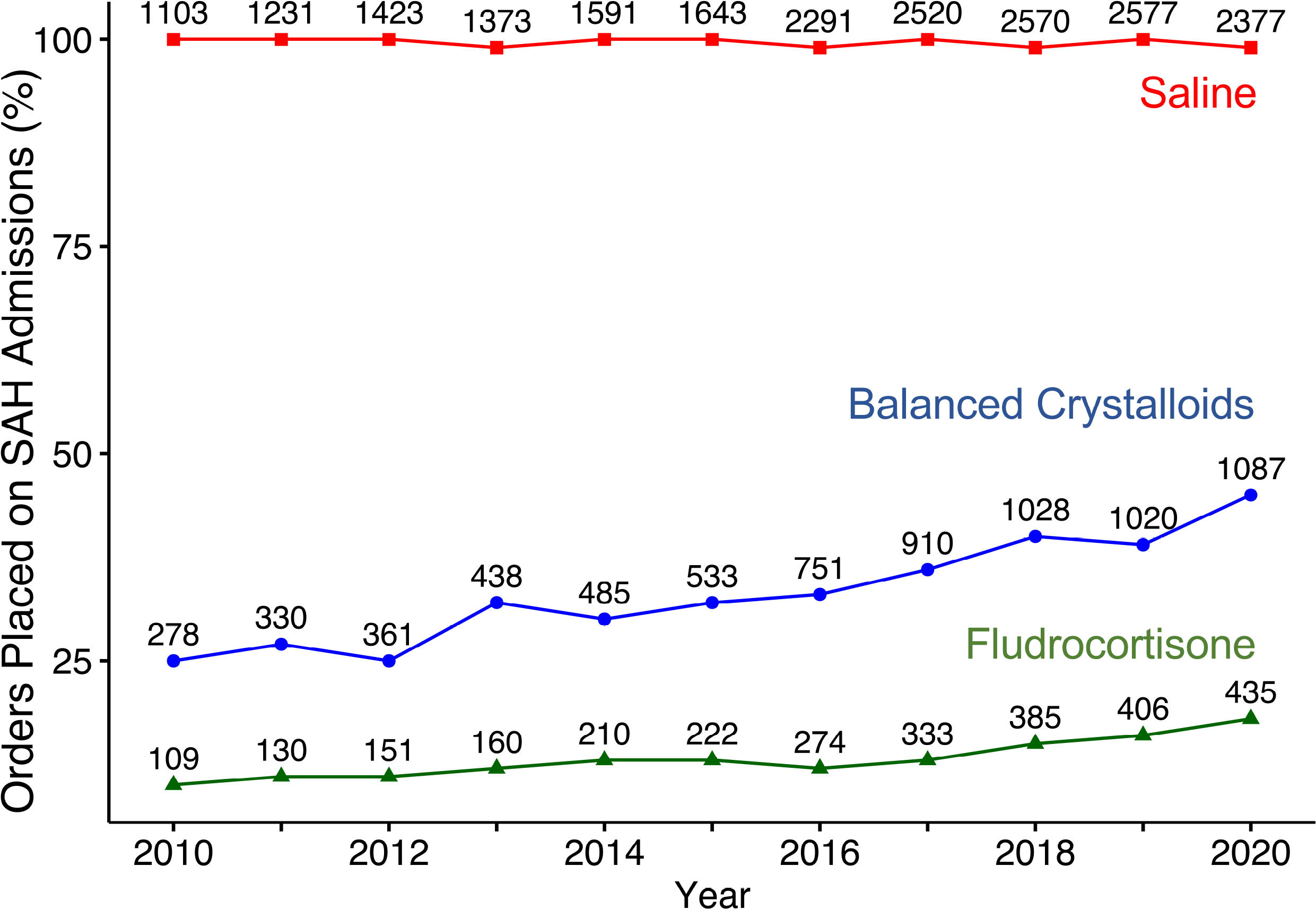
Fludrocortisone and crystalloids usage rates in subarachnoid hemorrhage patients in the Premier Healthcare Database. From 2010 to 2020, 20,699 subarachnoid hemorrhage encounters were retrieved across about 350 hospitals in the United States with a CPT code involved in treating the ruptured aneurysm. The number above the dot indicates the actual number of encounters.

## DISCUSSION

Despite the recommendation by the 2023 (and in the previous 2012) American Heart Association aSAH guidelines that “the use of mineralocorticoids is reasonable to treat natriuresis and hyponatremia”(11) with a randomized level of evidence (Level B – Randomized) and moderate strength of evidence (2a – moderate; Benefit >> Risk), the use of mineralocorticoids in aSAH among the surveyed NINDS StrokeNet sites, which are largely tertiary academic centers was 70%. Even more noteworthy is its use is less than 25% in real-world practice data from Premier Healthcare Database. In the latter analysis, we did not include hydrocortisone which has the second highest but non-significant mineralocorticoid activity (relative potency of 0.008) compared to fludrocortisone (relative potency of 1)(13), because it is also commonly used to treat other conditions and may not accurately capture our aim to understand its use with the intent to treat natriuresis or hyponatremia. However, even with its inclusion, the usage rate of mineralocorticoids in aSAH patients is relatively lower in real-world practice compared to tertiary academic centers. Recent publications (14) suggest that uncertainty regarding the benefits of fludrocortisone treatment on aSAH outcomes may be a reason behind its low usage rates.

The usage rates of treatments of hyponatremia by the NINDS StrokeNet sites contrast those reported in the survey of 32 neurosurgical units in the United Kingdom and Ireland (15). In the latter survey, treatments used for hyponatremia were enteral salt supplementation (N=24, 75%), fluid restriction (N=18, 56.2%), hypertonic saline (N=17, 53.1%), and fludrocortisone (N=7, 21.9%). Collectively, data from the United States, United Kingdom, and Ireland sites not only demonstrate geographic variation in the treatment, but also commonly highlight the lack of a protocol in the sites to treat hyponatremia. Among the NINDS StrokeNet sites, only 24 (27.6%) sites have a protocol. Similarly, only 9 (28.1%) of the surveyed sites in the United Kingdom and Ireland have a protocol.

Maintaining intravascular volume is critical in aSAH patients, and crystalloid solutions are recommended and most commonly used. However, the formulation which offers the best outcomes for aSAH patients is unknown. Saline (0.9% NaCl) is the most commonly used crystalloid. Several randomized controlled trials in general and specific ICU patients have shown that balanced crystalloids, which have electrolyte composition similar to plasma, offer better outcomes compared to saline (16, 17). In our analysis, we estimated the usage rates of two balanced crystalloids—Plasma-Lyte or lactated Ringer’s—which are most commonly used. A limitation of our data from the Premier Healthcare Database is the inability to assess which was the primary fluid used since we captured the “order” for the fluid, not the quantity. Furthermore, there are other formulations such as Normosol-R, Isolyte S, Ringer’s acetate, and Hartmann’s solution whose usage rates we did not capture (18). Therefore, the collective usage rate of balanced crystalloid is probably slightly higher than what we report. However, in the context of increasing comparative effectiveness trials of balanced crystalloids versus saline, our data from the Premier Healthcare Database are noteworthy for the increasing use of balanced crystalloids in aSAH patients. Given their increase in usage, it is critical to study the impact of these crystalloid formulations in aSAH patients because a considerable volume of crystalloids (>16 liters during the ICU stay) is given to them to maintain euvolemia (8-10). The volume given in the randomized control trials pales in comparison. Meager evidence exists to inform crystalloid selection in aSAH patients; one small study shows worse outcomes in aSAH patients with the use of balanced crystalloids (8). Their increasing use in the United States is an important reason to study the impact of different formulations of crystalloids used for intravenous fluid therapy.

## CONCLUSIONS

Despite recommendations by the American Heart/Stroke Association guidelines, the use of mineralocorticoids remains low, and balanced crystalloids are increasingly used as fluid therapy in aSAH patients. If mineralocorticoids and balanced crystalloids improve outcomes of aSAH patients must be rigorously tested.

## Data Availability

All data produced in the present study are available upon reasonable request to the authors

## Notes

### Competing Interest Statement

The authors have declared no competing interest.

### Funding Statement

This study was funded by NIH/NINDS U01NS086872 and U01NS087748

### Author Declarations

We analyzed the service-level, all-payer Premier Healthcare Database (PINC AI / Premier Inc., Charlotte, NC) of electronic health records and hospital billing data, and a survey of institution-based clinical practice was performed, rather than individual physician-based clinical practice.

## REFERENCES

1. Chan V, Lindsay P, McQuiggan J, et al. Declining Admission and Mortality Rates for Subarachnoid Hemorrhage in Canada Between 2004 and 2015. Stroke 2019;50:181–184.

2. Lee VH, Ouyang B, John S, et al. Risk stratification for the in-hospital mortality in subarachnoid hemorrhage: the HAIR score. Neurocrit Care 2014;21:14–19.

3. Stienen MN, Germans M, Burkhardt JK, et al. Predictors of In-Hospital Death After Aneurysmal Subarachnoid Hemorrhage: Analysis of a Nationwide Database (Swiss SOS [Swiss Study on Aneurysmal Subarachnoid Hemorrhage]). Stroke 2018;49:333–340.

4. Nieuwkamp DJ, Setz LE, Algra A, et al. Changes in case fatality of aneurysmal subarachnoid haemorrhage over time, according to age, sex, and region: a meta-analysis. Lancet Neurol 2009;8:635–642.

5. Chen S, Li Q, Wu H, et al. The harmful effects of subarachnoid hemorrhage on extracerebral organs. Biomed Res Int 2014;2014:858496.

6. Lantigua H, Ortega-Gutierrez S, Schmidt JM, et al. Subarachnoid hemorrhage: who dies, and why? Crit Care 2015;19:309.

7. Solenski NJ, Haley EC, Jr., Kassell NF, et al. Medical complications of aneurysmal subarachnoid hemorrhage: a report of the multicenter, cooperative aneurysm study. Participants of the Multicenter Cooperative Aneurysm Study. Crit Care Med 1995;23:1007–1017.

8. Mistry AM, Magarik JA, Feldman MJ, et al. Saline versus Balanced Crystalloids for Adults with Aneurysmal Subarachnoid Hemorrhage: A Subgroup Analysis of the SMART Trial. Stroke Vasc Interv Neurol 2022;2.

9. Mistry AM, Mistry EA, Ganesh Kumar N, et al. Corticosteroids in the Management of Hyponatremia, Hypovolemia, and Vasospasm in Subarachnoid Hemorrhage: A Meta-Analysis. Cerebrovasc Dis 2016;42:263–271.

10. Barlow B, Thompson Bastin ML, Shadler A, et al. Association of chloride-rich fluids and medication diluents on the incidence of hyperchloremia and clinical consequences in aneurysmal subarachnoid hemorrhage. J Neurocrit Care 2022;15:113–121.

11. Hoh BL, Ko NU, Amin-Hanjani S, et al. 2023 Guideline for the Management of Patients With Aneurysmal Subarachnoid Hemorrhage: A Guideline From the American Heart Association/American Stroke Association. Stroke 2023;54:e314–e370.

12. Oddo M, Poole D, Helbok R, et al. Fluid therapy in neurointensive care patients: ESICM consensus and clinical practice recommendations. Intensive Care Med 2018;44:449–463.

13. Schimmer BP, Funder JW. Adrenocorticotropic Hormone, Adrenal Steroids, and the Adrenal Cortex. In: Brunton LL, Hilal-Dandan R, Knollmann BC, eds. Goodman & Gilman’s: The Pharmacological Basis of Therapeutics, 13e. New York, NY: McGraw-Hill Education, 2017.

14. Busl KM, Rabinstein AA. Prevention and Correction of Dysnatremia After Aneurysmal Subarachnoid Hemorrhage. Neurocrit Care 2023.

15. Tominey S, Baweja K, Woodfield J, et al. Investigation and management of serum sodium after subarachnoid haemorrhage (SaSH): a survey of practice in the United Kingdom and Republic of Ireland. Br J Neurosurg 2021:1–4.

16. Collins MG, Fahim MA, Pascoe EM, et al. Balanced crystalloid solution versus saline in deceased donor kidney transplantation (BEST-Fluids): a pragmatic, double-blind, randomised, controlled trial. Lancet 2023;402:105–117.

17. Semler MW, Self WH, Wanderer JP, et al. Balanced Crystalloids versus Saline in Critically Ill Adults. N Engl J Med 2018;378:829–839.

18. Semler MW, Kellum JA. Balanced Crystalloid Solutions. Am J Respir Crit Care Med 2019;199:952–960.

